# Examining the decision to offer in-person college instruction during the COVID-19 era: A multilevel analysis of the factors that affected intentions to open

**DOI:** 10.1101/2020.10.15.20213363

**Authors:** Jacob Louise Felson, Amy Adamczyk

**Affiliations:** William Paterson University; John Jay College of Criminal Justice and The Graduate Center, City University of New York

## Abstract

At the end of Summer 2020 colleges and universities had to make difficult decisions about whether to return to in-person instruction. While opening campuses could pose a major health risk, keeping instruction online could dissuade students from enrolling. Taking an ecological approach that considers the influence of state, county, and college characteristics, this study uses mixed modeling techniques and data from 89% of two- and four-year public and four-year private US colleges to assess the factors that shaped their decision to provide mostly in-person instruction as of August 1, 2020. We consider the roles of the political and religious climate, COVID-19 infections, deaths, and mask mandates, college niche, finances, dormitory capacity, faculty resistance, online readiness, and enrollment pressures. Most notably, we find that decision-making was unrelated to cumulative COVID infection and related mortality rates. The strongest predictor of in-person instruction was the proportion of state residents who voted for Trump in the 2016 presidential election. We also find that dormitory capacity, percentage of revenue from tuition, institutional importance to the local economy, graduation rates, and per capita endowment were associated with providing in-person instruction.

---

Beginning in March 2020, COVID-19 became a major impediment to face-to-face businesses. The virus was spreading rapidly and state governments began issuing stay-at-home orders to workers deemed non-essential (Mervosh, Lu, and Swales 2020). Colleges across the country transitioned from in-person to online instruction, resulting in many students returning to their parents’ homes to complete their Spring semester online (Marsicano et al. 2020). Gradually, COVID-19 infections and deaths began to decline and by summer many states that had previously closed were now open. Summer gatherings resulted in a COVID-19 infection surge in many parts of the country that had not previously been affected (Foster and Mundell 2020). This latter surge was apparently concentrated among younger people and thus resulted in less serious illness and fewer deaths (Colson 2020).

In deciding whether to resume in-person classes, college administrators faced significant cross-pressures. On one hand, colleges faced financial and political pressure to reopen. Administrators at many schools could expect steep enrollment declines if they kept instruction online since distance learning is generally perceived as a poor substitute. Politically, President Trump and his supporters were especially vocal in their support of colleges fully reopening (Bauer-Wolf 2020). On the other hand, colleges faced pressure from many faculty and some health officials to remain online. Of particular concern was that the traditional college-age population was being infected at higher rates and would not be as likely to heed public health warnings about social distancing and mask wearing (Anon 2020; Wan and Balingit 2020). Confronted by conflicting pressures, many colleges waited until August to make decisions regarding in-person instruction.

Drawing on a sample of 89% of America’s non-specialized two- and four-year public and private colleges,^1^ our study assesses the factors that shaped reopening decisions as of August 1, 2020. We chose to focus on tentative plans in mid-summer rather than on final choices since we seek to understand administrative decision-making at a time when they conceivably had maximal options. As the summer progressed, alternatives for some administrators narrowed due to state health rules and COVID-19 outbreaks.

Drawing on a Weberian understanding of rationality and bureaucracy (Weber 1978 [1921]; Waters and Waters 2015), we view school administrators as engaging in rational cost-benefit calculations. Using multilevel modeling techniques and data drawn from a range sources, we consider a total of twenty-eight factors at the state, county, and institutional levels. Though there are nuances in the conclusions that we discuss below, the bottom-line is that, to the extent decision-making could be predicted, it was driven by state politics, enrollment pressure and product niche, not by local COVID-19 infection or related fatality rates.

## UNIVERSITY DECISION MAKING

To understand college administrators’ decisions, we draw on a Weberian perspective of bureaucracy and rationality (Weber 1978 [1921]; Waters and Waters 2015). Within this framework, decision-making is viewed as grounded within a process of bounded rationality (Simon 1990). For college administrators the decision-making process entails choosing from among alternatives to achieve a certain result (Eisenfuhr 2011). In deciding to reopen in-person instruction college executives had to consider whether it was feasible, what the alternatives might be and what impact it would have (Grant 2011). Many people would be impacted by the decision to reopen, including the campus community and nearby residents. Likewise, administrators had to make virtually unprecedented decisions within complex economic, political, health, and cultural environments with limited knowledge.

Many college administrators’ decisions had to be made jointly with leaders at the system- and state levels. As we investigate the factors impacting reopening, we draw on an ecological perspective (Duncan, Schnore, and Rossi 1959) that considers not only the characteristics of colleges, but also the dynamics operating in the geographical context and accounts for system-level influences. As we explain in the next section, the norms, political preferences, laws, and experiences with COVID-19 in the surrounding area were likely to play a role in deciding to reopen.^2^

## LOCAL AREA CHARACTERISTICS

### Political partisanship

One of the most important factors to consider for understanding the likelihood that colleges would open campuses, is support for Trump in the 2016 presidential election. President Trump had encouraged schools of all kinds to resume in-person instruction in the fall (Bauer-Wolf 2020). He chose to support the requirement that international students could not take more than one on-line course each term, which would have forced international students at colleges with all online instruction to leave the country (Schwartz 2020). Several other political leaders have followed Trump’s lead in minimizing the extent of the pandemic, its detrimental consequences, and encouraging businesses to reopen (Olorunnipa, Witte, and Bernstein 2020).

Part of college administrators’ jobs is to look after the political and economic interests of their institutions, and those interests vary at the state and local levels. Thus, we would expect that support for Trump within counties and states would impact reopening decisions. In addition to different cultural and economic effects, policy makers’ interests are based on where their constituents are located (Saffell and Basehart 1997). As the proportion of residents who voted for Trump in the 2016 election in either states or counties increases, norms and policies should be more likely to reflect pro-Trump sentiments, exerting pressure on colleges to provide in-person instruction.

Additionally, since they receive part of their funding from the state government, the greatest effect of the proportion of residents who voted for Trump should be felt by public rather than private colleges. Figure 1 presents a map of the United States with bluer colors indicating that a greater proportion of people voted for Hillary Clinton in the 2016 election and redder colors showing less support. For the three largest public four-year colleges in each state, yellow dots indicate that they had chosen to provide mostly online instruction or were undecided as of August 1, 2020. Green dotes note that they were planning for regular in-person instruction. Some of the bluest states are California, Washington, Oregon, and Illinois, where only two of the 12 largest four-year public colleges were planning to open (i.e., green dots) their campuses.

**Figure 1:**
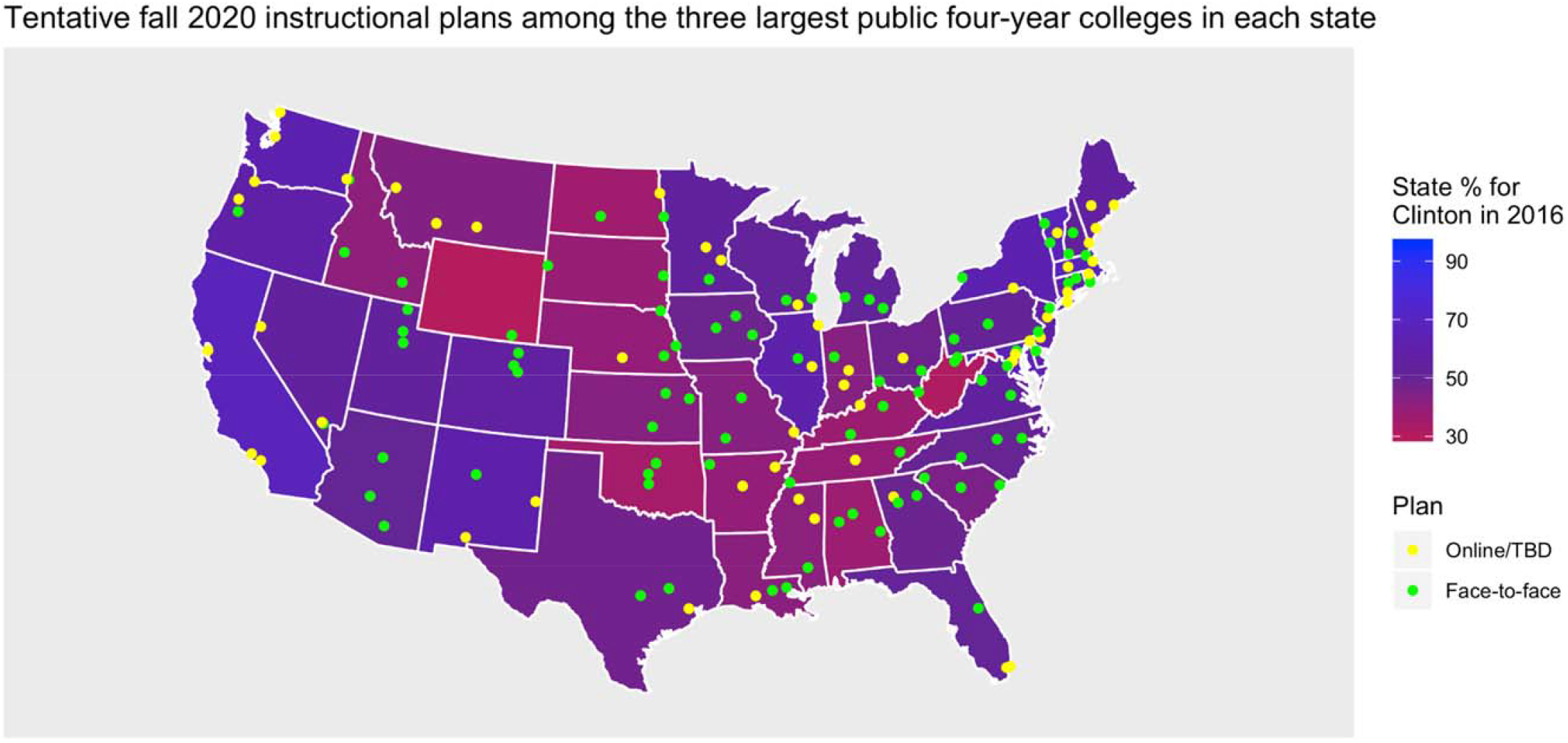
**Note**: To make the map easier to see, only the three largest public four-year colleges are included for each state. Note the increase in online instruction (i.e., yellow dotes) in bluer states, like California, Washington, Oregon, and Illinois.

### Concentrations of conservative Protestants

Early in the COVID-19 crisis, several news stories reported that leaders of some conservative churches across the United States were downplaying the severity of the pandemic and were continuing to gather in-person despite stay at home orders (Barria 2020; Kaleem 2020). Evangelicals’ greater reluctance to heed warnings from public health authorities could have followed from Trump’s example but could also have roots in the greater skepticism toward scientific institutions observed among religious people (Evans 2013; Gauchat 2008; Hill, Gonzalez, and Burdette 2020). Areas with higher concentrations of Evangelicals may be more inclined to support norms that do not take COVID-19 as seriously as people in other parts of the country. College administrators may be affected by local conservative religious sentiments, resulting in them being more likely to offer in-person classes.

### Colleges’ economic impact on the surrounding area

Colleges that make a major contribution to the local economy may also be under greater pressure to provide in-person instruction (Sullivan 2020). In communities where the college is relatively large compared to the local population, businesses (e.g., souvenir shops, food vendors) are likely to have arisen to meet the demands of students, employees, parents, and others (Grady 2017). They are also likely to employ local residents to work on campus (Gumprecht 2003). In deciding whether or not to allow for in-person classes, colleges may consider the economic and cultural impact of their decision on the local community, especially in places where they are a major source of revenue.

### Local COVID-19 infections and fatalities

We would expect that administrators of colleges in states and counties with higher cumulative rates of COVID-19 infections and fatalities to be less likely to plan for in-person classes. The more proximate the suffering from the virus is, the more tangible and concerning it would likely be for college leaders and the rest of the campus community alike. Thus, we would expect administrators in highly affected areas to have more personal, political and financial reasons to remain remote in the fall. Administrators in such areas might reasonably conclude that parents in communities ravaged by COVID-19 might not want their children returning to in-person instruction anyway, and so the risk to enrollment of remaining online might be minimal. The political pressure to remain remote might also be greater in areas where death rates were high.

## INSTITUTIONAL CHARACTERISTICS

While local and state cultures, political climates, economics, and infection rates are likely to have a role in shaping reopening decisions, the characteristics of colleges themselves should have a major impact. In our study we consider the role of five sets of college characteristics -- product niche, finances, online readiness, faculty support, and enrollment pressure-that could shape the odds that they offer mostly in-person classes in early August, 2020.

### Product niche

Brick-and-mortar colleges vary dramatically in the kinds of educational products they offer and in the extent to which remote learning offers a viable substitute. Two-year community colleges and lower-cost four-year public schools offer commodity educational products that compete largely on price within their local markets. By contrast, higher cost four-year public and private schools compete less on cost and more on the quality of the educational goods they purport to provide (Sun 2020). Students attending the highest-cost private schools are paying for what amounts to an exclusive club -- bespoke treatment, ample campus amenities and access to networking opportunities (Holmstrom, Karp, and Gray 2011). This exclusive experience is of course highly dependent on face-to-face interaction.

The more exclusive the education, the less that distancing learning is likely perceived as a viable substitute by students or administrators seeking to serve their institutional mission and keep students happy. And institutions offering high-priced degrees are limited in their ability to reduce prices and product offerings due to high fixed costs and concerns about maintaining their brand.

Schools that spend more per student and are more selective should also be more likely to plan for in-person instruction because they will sacrifice more than others by switching to online instruction. In particular, the networking benefits of attending a selective school are likely to be attenuated significantly by keeping instruction remote.

We also expect that schools with larger proportions of students living on campus to be more likely to reopen. In the United States, the residential college living experience is often seen as a rite of passage. Colleges generate a lot of revenue from having students live on campus, where they not only pay for a room, but also the accompanying meal plan and often extracurricular activities (Lederman 2020). Likewise, colleges with a higher proportion of students residing on campus are going to have student bodies that are expecting the full residential college experience. For students planning to attend college while living with parents or elsewhere off-campus, online courses may be less of a letdown, and in some cases online delivery may make attending classes easier, eliminating the commute (Castonguay 2020).

### Faculty resistance: Faculty unions and percentage full professor

Often faculty members and their unions have strong preferences, which colleges may have considered in making their decision. Many universities have a tenure track system whereby faculty move through the ranks of assistant, associate and full professor. Following eight or more years of education, it can take a lot of time before faculty reach the rank of full professor. As a result, faculty tend to be older than the general working population. While just 23% of people in the general workforce are over the age of 55, 37% of faculty are (McChesney and Bichsel 2020). Older individuals are especially vulnerable to severe complications related to the coronavirus (CDC 2020b). When a university has a higher proportion of full professors, the faculty may advocate for more on-line teaching to protect their health. Likewise, a substantial number of colleges have a faculty union, which may advocate on behalf of faculty and staff to resist in-person classes (Cain 2017). Faculty pressure may therefore play a role in shaping a college’s decision to offer in-person classes.

### Finances

A college’s financial well-being is also likely to shape their willingness to provide in-person instruction, though less clear is the direction of the relationship. Financially vulnerable schools may be more likely to provide in-person classes to retain students who do not want to take online courses (Lederman 2020). In making a reopening decision, colleges have to consider the possibility that students may go elsewhere, threatening the school’s financial sustainability (Quintana 2020). Similarly, when colleges have larger endowments, they may be able to take the revenue hit related to some students temporarily or permanently withdrawing.

It is also possible that financial well-being is related to an increased likelihood of in-person classes. In order for in-person instruction to occur, colleges need to enact a range of safety measures (e.g., virus testing, personal protective equipment, temperature checks, ventilation systems) which can be costly (ACE 2020; CDC 2020a). Schools that are more financially secure (i.e., higher net revenue and endowment per student) may have the financial resources to open safely enough to convince students (and their parents) to return, thereby further increasing their financial security. Additionally, these colleges may be more likely to offer services, activities, and events, like college football games, that can provide a lot of revenue, further improving their financial position. Colleges in a worse financial state may be less likely to historically offer these revenue generating activities, and hence would suffer less by moving most or all classes online.

### Online readiness

The final characteristic that our study considers is the extent to which colleges are prepared to teaching online. Before COVID-19 shut down most in-person higher learning in March 2020, colleges varied substantially in the extent to which they were offering online classes. While about 20% of students at public schools have had some online instruction, about 9% of nonprofit private schools have (Lederman 2018). The more that colleges were offering online classes before the pandemic, the more skilled in providing remote learning they should be. Likewise, schools that are providing more virtual teaching should also be more likely to have student bodies that are familiar with and less averse to this alternative form of instruction.

In the next section we explain how we assess which factors are most likely to shape the odds of having in-person classes. One of the strengths of our study is that we will be able to see which characteristics overlap and isolate those that have a unique effect in explaining a college’s decision to offer all in-person instruction in early August 2020. Our analysis will not only examine college characteristics, but also the multilevel influences of the state and local political climate and cultural norms. At the time of submission, this is the only peer reviewed study^3^ that we know of that comprehensively examines the factors shaping college reopening plans.

## Data and Methods

Descriptions and sources of data are presented in Table 1. For planned Fall 2020 instruction modality, we relied on data from Davidson’s College Crisis Initiative (CCI) (2020), which includes information on reopening plans as of August 1, 2020 for approximately 89% of US institutions of higher education offering two- and four-degrees in multiple subject areas. This information was retrieved from the Chronicle of Higher Education (2020). Coverage of more specialized institutions and tribal colleges in the CCI database was meager (33%), so we excluded those institutions from our analysis. Our analytic sample included the vast majority of institutions with 2018 Carnegie classifications of associate’s college (88%), associate/baccalaureate and baccalaureate college (83%), master’s institution (90%) and doctoral university (95%). Overall, the sample included 2,664 (approximately 89%) of all such institutions listed in the 2018 Integrated Postsecondary Education Data System database.

**Table 1:**
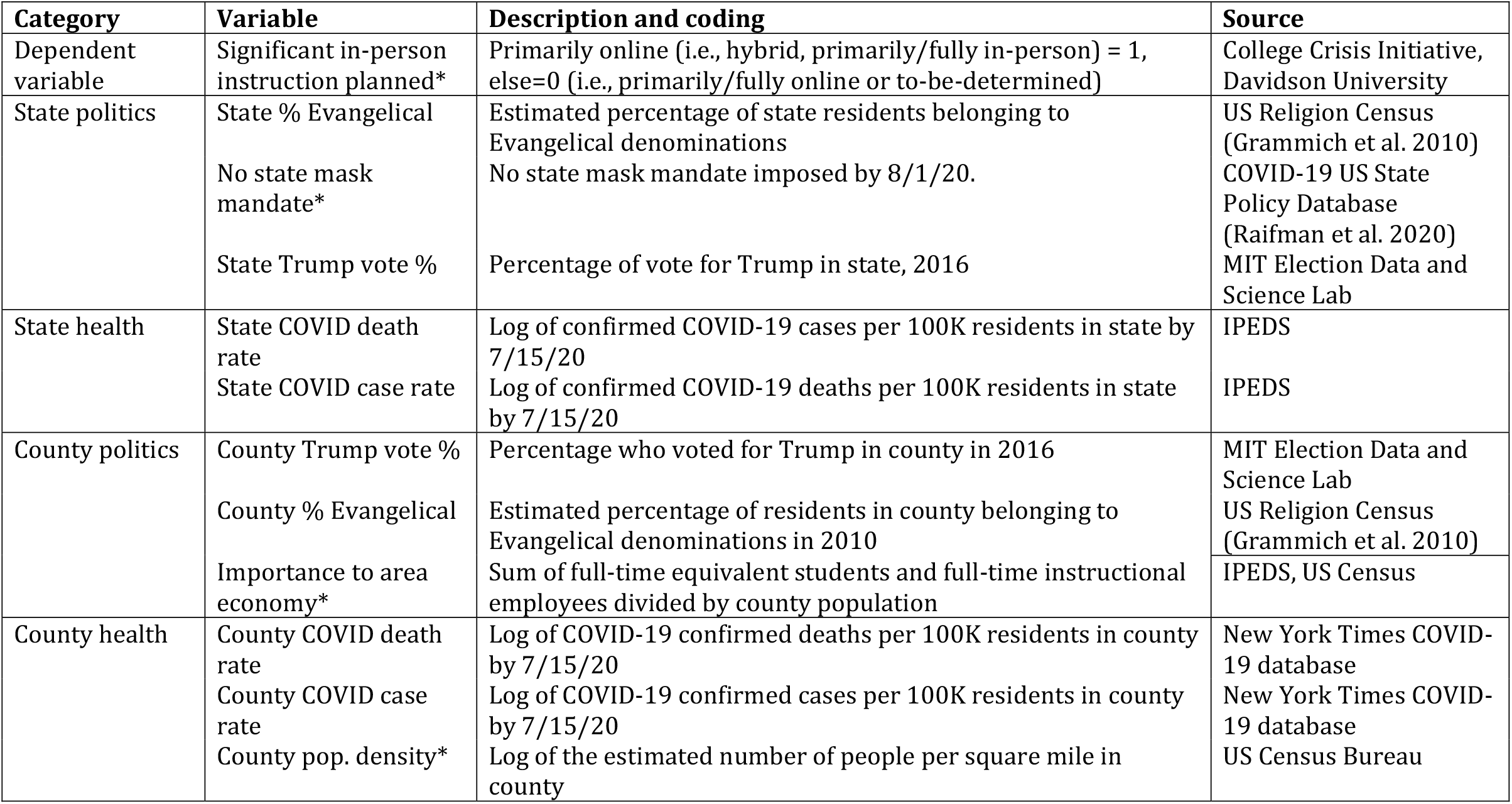
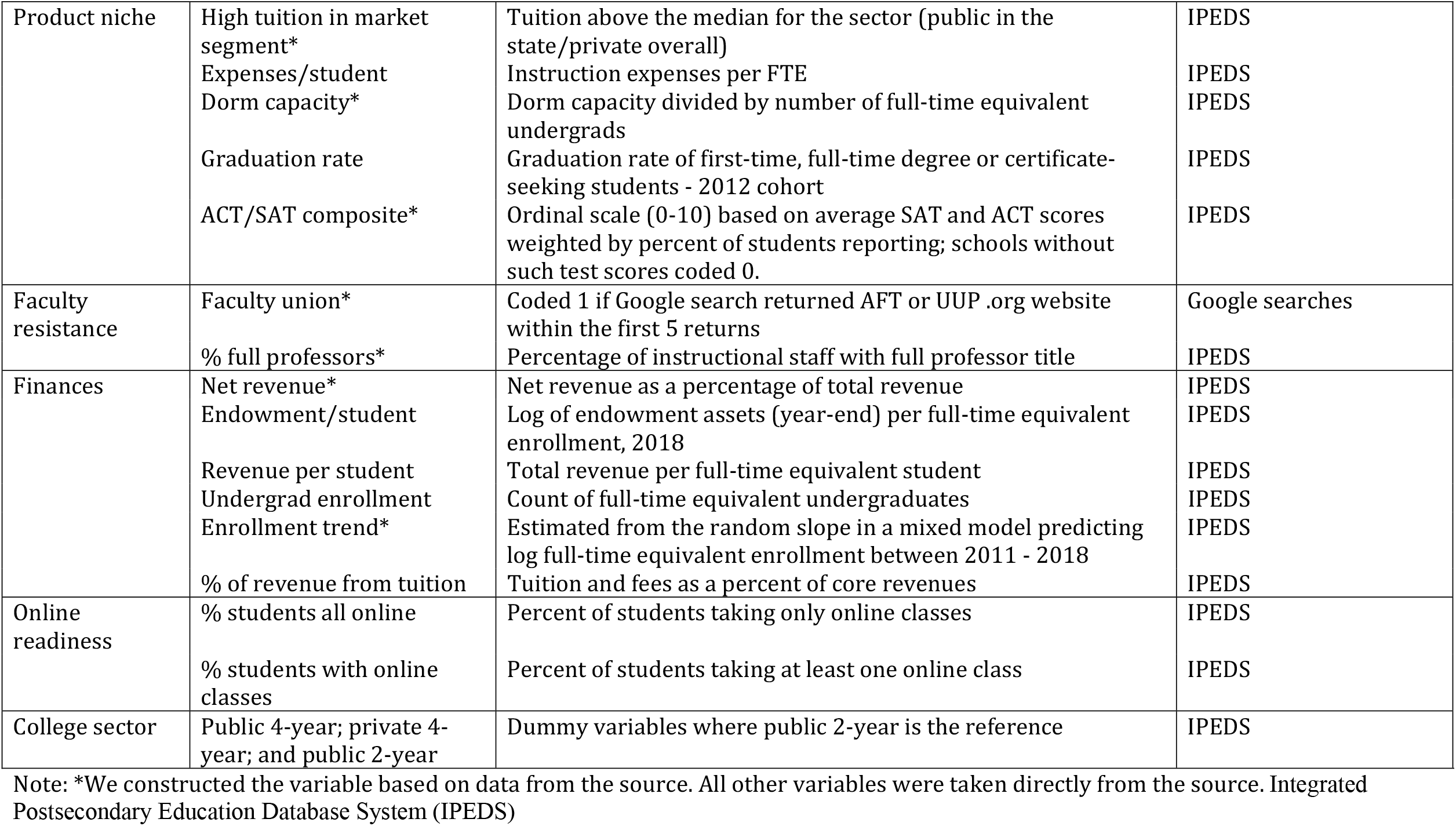
Description and sources of variables in the analysis

CCI had classified fall instructional plans into one of six categories: to-be-determined, fully online, primarily online, hybrid, primarily in-person, and fully in-person. We take the former three categories (i.e., to-be-determined, fully online, primarily) to be indicative of a more cautious approach of returning to classes. The latter three categories (i.e., hybrid, primarily in-person, and fully in-person) by contrast indicate a decision to return substantial numbers of students and faculty to the classroom in the face of considerable uncertainty about the associated risks.

As shown in Table 1, most of our independent variables come from the Integrated Postsecondary Education Database System (IPEDS) based at the National Center for Education Statistics. The coding of most variables was straightforward, but several coding decisions require further explanation. We measure tuition and fees (in 2018) as a dichotomous variable indicating whether it fell above the median for the state among public institutions, and whether it fell above the median for the country among private institutions. The idea here is to (1) gauge whether the institution is charging a premium in the market where it operates using (2) a measure that is not collinear with institutional sector (public/private), which tends to be highly correlated with the cost of tuition.

Using these data, we estimated hierarchical logistic regression models predicting whether or not college administrators had announced plans for significant classroom instruction in their institution for the fall semester by August 1, 2020. Our models include random effects for states as administrator decision-making is assumed to correlate within them. Within states, we found minimal evidence for spatial autocorrelation using Moran’s I on the original five-category ordinal measure from CCI. Thus, we conclude that decision-making is unrelated to geographic proximity within states.

Our models also include random effects for systems of higher education within which 41% of institutions in our sample are subsumed. Since only a subset of institutions is subsumed within systems of higher education, our data can be described as partially nested, which complicates modeling decisions. There is no canonical modeling strategy for partially nested data with binary outcomes. Research in this area has focused on evaluating methods for estimating the effects of treatments administered to individuals nested within clusters (e.g. classes, therapists) when individuals in the control group are not nested (Roberts, Batistatou and Roberts, 2016; Lohr, Schochet and Sanders, 2014). The methodological literature in the area is limited in its ability to inform our study in that we are not comparing outcomes between nested and non-nested cases.

We estimated two sets of models in accordance with alternative strategies presented in the literature cited above. Both sets of models include a random intercept for state but differ in how they handle nesting within systems of higher education (e.g. California State, Pennsylvania State, etc.). One set of models also includes a random effect of a dummy variable indicating affiliation with a larger system that varies by system. The other set of models includes a random intercept varying by system, where unaffiliated colleges are all included in one pseudo-cluster.^4^ Since the results from both sets of models are substantively similar, we present results from only the former.

In our models, we standardized all continuous (non-dichotomous) independent variables to facilitate interpretation. We logged continuous variables with absolute skewness values above 1.5, reasoning that non-linear effects were generally more plausible in those cases.

Model-building went as follows. First, we estimated a Model for each of the eight themes of factors which we had reason to believe would influence the decision to return to face-to-face teaching. Then, we estimated a final Model that included all variables for which there were significant coefficients in the thematic models.

## Results

Table 2 presents descriptive statistics for all variables in the analysis prior to standardization. Table 3 presents averages of each of the unstandardized independent variables separately by college plan. The label of the rightmost column of Table 3, TBD, refers to institutions in which administrations had announced that teaching modalities were yet to be determined.

**Table 2:** Descriptive statistics for unstandardized versions of variables included in the analysis

|  | Mean | SD | Min | Max |
| --- | --- | --- | --- | --- |
| Significant in-person instruction planned | 0.44 | 0.50 | 0.00 | 1.00 |
| State % Evangelical | 16.89 | 10.57 | 2.28 | 42.04 |
| No state mask mandate | 0.22 | 0.42 | 0.00 | 1.00 |
| State Trump vote % | 47.62 | 9.75 | 4.12 | 70.05 |
| 4-year public | 0.28 | 0.45 | 0.00 | 1.00 |
| 4-year private | 0.37 | 0.48 | 0.00 | 1.00 |
| County Trump vote % | 48.31 | 17.47 | 4.12 | 87.81 |
| County % Evangelical | 17.80 | 13.41 | 0.05 | 73.00 |
| Importance to area economy | 0.27 | 1.53 | -6.46 | 4.28 |
| County COVID death rate | 2.80 | 1.43 | 0.00 | 5.91 |
| County COVID case rate | 6.52 | 0.92 | 2.83 | 9.09 |
| County pop. density | 5.86 | 1.74 | 0.91 | 11.18 |
| State COVID death rate | 3.42 | 1.11 | 0.00 | 6.47 |
| State COVID case rate | 6.88 | 0.67 | 4.51 | 8.79 |
| Faculty union | 0.30 | 0.46 | 0.00 | 1.00 |
| % full professors | 21.37 | 18.01 | 0.00 | 100.00 |
| Net revenue | 0.05 | 0.15 | -1.00 | 0.79 |
| Endowment/student | 7.77 | 3.43 | 0.00 | 14.93 |
| Revenue per student | 10.03 | 0.53 | 8.69 | 13.07 |
| Enrollment trend | -0.01 | 0.04 | -0.32 | 0.48 |
| Undergrad enrollment | 7.95 | 1.08 | 3.87 | 10.92 |
| % of revenue from tuition | 38.59 | 25.27 | 0.00 | 100.00 |
| % students all online | 10.98 | 10.70 | 0.00 | 50.00 |
| % students with online classes | 18.98 | 14.42 | 0.00 | 100.00 |
| High tuition in market segment | 0.51 | 0.50 | 0.00 | 1.00 |
| Expenses/student | 8.99 | 0.50 | 7.19 | 11.76 |
| Dorm capacity | 2.24 | 1.94 | 0.00 | 4.62 |
| Graduation rate | 45.62 | 20.74 | 0.00 | 100.00 |
| ACT/SAT composite | 2.69 | 3.41 | 0.00 | 10.00 |

**Table 3:** Average of each unstandardized independent variable by college’s fall plan on August 1, 2020

|  | Fully/primarily<br>online | TBD | Hybrid | Primarily/fully<br>in-person |
| --- | --- | --- | --- | --- |
| State % Evangelical | 15 | 19 | 17 | 18 |
| No state mask mandate | 0.16 | 0.26 | 0.17 | 0.30 |
| State Trump vote % | 45 | 48 | 48 | 50 |
| 4-year public | 0.29 | 0.20 | 0.37 | 0.28 |
| 4-year private | 0.21 | 0.32 | 0.45 | 0.54 |
| County Trump vote % | 44 | 51 | 48 | 52 |
| County % Evangelical | 15 | 20 | 17 | 20 |
| Importance to area econ. | 0.16 | 0.11 | 0.24 | 0.61 |
| County COVID death rate | 2.9 | 2.7 | 3.0 | 2.6 |
| County COVID case rate | 6.6 | 6.5 | 6.6 | 6.4 |
| County pop. density | 6.1 | 5.6 | 6.0 | 5.7 |
| State COVID death rate | 3.5 | 3.4 | 3.6 | 3.2 |
| State COVID case rate | 6.9 | 6.9 | 7.0 | 6.8 |
| Faculty union | 0.39 | 0.26 | 0.33 | 0.22 |
| % full professors | 20 | 18 | 23 | 24 |
| Net revenue | 0.046 | 0.026 | 0.078 | 0.050 |
| Endowment/student | 6.9 | 6.7 | 8.7 | 9.1 |
| Revenue per student | 10.0 | 9.9 | 10.0 | 10.0 |
| Enrollment trend | -0.0120 | -0.0180 | -0.0097 | -0.0065 |
| Undergrad enrollment | 8.3 | 7.6 | 8.0 | 7.9 |
| % of revenue from tuition | 30 | 35 | 44 | 47 |
| % students all online | 12.0 | 12.0 | 9.4 | 11.0 |
| % students w online classes | 20 | 21 | 19 | 16 |
| High tuition in market segment | 0.49 | 0.35 | 0.66 | 0.59 |
| Expenses/student | 9.0 | 8.9 | 9.1 | 9.1 |
| Dorm capacity | 1.5 | 1.7 | 2.9 | 3.1 |
| Graduation rate | 42 | 39 | 52 | 52 |
| ACT/SAT composite | 2.2 | 1.5 | 3.8 | 3.6 |

Note: To-be-determined (TBD)

Table 4 shows coefficients for each of the independent variables when included separately in hierarchical logistic regressions (specified as described above) predicting significant in-person instruction. Significant coefficients in the expected direction are bolded; significant coefficients in the opposite direction expected are italicized. Since all continuous variables are standardized, we can compare their magnitudes. We can roughly compare the magnitudes of dichotomous variables with those of continuous variables by multiplying the coefficients of the latter by two, capturing a change from −1 SD to + 1 SD.

**Table 4:** Effects of variables in bivariate logistic regressions with random effects for examining mostly (i.e., hybrid, primarily/fully online) in-person instruction. (Exponentiated odds coefficients shown)

| Predictor | beta | Predictor | beta |
| --- | --- | --- | --- |
| State % Evangelical | 1.06 | % full professors | 1.19*** |
| No state mask mandate | 0.98 | Net revenue | 1.11* |
| State Trump vote % | 1.21* | Endowment/student | 2.13*** |
| 4-year public | 3.68*** | Revenue per student | 1.25*** |
| 4-year private | 4.85*** | Enrollment trend | 1.14* |
| County Trump vote % | 1.09 | Undergrad enrollment | 1.09+ |
| County % Evangelical | 1.1 | % of revenue from tuition | 1.52*** |
| Import to area economy | 1.21*** | % students all online | 0.87** |
| County COVID death rate | 0.95 | % students with online classes | 0.8*** |
| County COVID case rate | 0.97 | High tuition in market segment | 1.47*** |
| County pop. density | 0.97 | Expenses/student | 1.42*** |
| State COVID death rate | 0.92 | Dorm capacity | 2.12*** |
| State COVID case rate | 0.98 | Graduation rate | 1.89*** |
| Faculty union | 0.94 | ACT/SAT composite | 1.57*** |

+p<0.01; \*p<0.05; \*\*p<0.01; \*\*\*p<0.001

We see that the variables most strongly associated with returning to campus are sector (i.e., four-year-private and four-year public school versus two-year institutions), endowment per student, dorm capacity and graduation rate. There are somewhat weaker associations with selectivity, percent revenue from tuition, high tuition in market segment and expenses per student. Changes in all of these variables (either 0 to 1 or −1 SD to + 1 SD) are associated with at least a doubling in the odds of planning face-to-face instruction. By contrast, county political and health variables are essentially unrelated to reopening plans.

Moving now to Table 5, Models 1 and 2 test state politics and the religious environment with and without interactions between 4-year public and private schools and state Trump vote. In Model 2 of Table 5, the effect of the proportion of state residents who voted for Trump on in-person planning is significant and substantial only among public schools. A post-hoc test shows that the 4-yr private institution x State Trump vote % interaction essentially cancels out the State Trump vote % effect (χ^2^ = 2.84; p=0.092). Models 3-5 in Table 5 echo the conclusions of the bivariate results, most notably that COVID rates are not significantly related to colleges’ decisions at either the state or county levels, and county political and religious environments are also irrelevant.

**Table 5:** Hierarchical (two-level) logistic models examining August 1, 2020 college plans to have any in-person instruction in the fall 2020 semester

|  | State politics |  | State health | County politics | County health |
| --- | --- | --- | --- | --- | --- |
|  | (1) | (2) | (3) | (4) | (5) |
| State % Evangelical | 0.84 | 0.84 |  |  |  |
| No state mask mandate | 0.75 | 0.72 |  |  |  |
| State Trump vote % | 1.54*** | 2.09*** |  |  |  |
| 4-year public | 3.73*** | 3.81*** |  |  |  |
| 4-year private | 4.98*** | 5.54*** |  |  |  |
| 4-yr private x Trump vote |  | 0.58*** |  |  |  |
| 4-yr public x Trump vote |  | 1.23 |  |  |  |
| State COVID death rate |  |  | 1.10 |  |  |
| State COVID case rate |  |  | 0.87 |  |  |
| County Trump vote % |  |  |  | 1.00 |  |
| County % Evangelical |  |  |  | 1.07 |  |
| Importance to area economy |  |  |  | 1.20** |  |
| County COVID death rate |  |  |  |  | 0.94 |
| County COVID case rate |  |  |  |  | 1.02 |
| County pop. density |  |  |  |  | 0.98 |
| Constant | 0.34*** | 0.31*** | 1.10 | 1.10 | 1.11 |
| State error SD | 0.39 | 0.41 | 0.52 | 0.50 | 0.52 |
| System error SD | 1.34 | 1.29 | 1.87 | 1.87 | 1.87 |
| Observations | 2,351 | 2,351 | 2,351 | 2,351 | 2,351 |
| Log Likelihood | -1,384.79 | -1,370.47 | -1,458.74 | -1,453.18 | -1,458.71 |
| Bayesian Inf. Crit. | 2,831.68 | 2,818.56 | 2,956.28 | 2,952.95 | 2,964.01 |

+p<0.01; p<0.05; p<0.01; p<0.001.
Note: All quantitative variables are standardized. The reference category for sector is two-year schools. The following variables are logged: endowment, state COVID deaths, state COVID cases, county COVID deaths, county COVID cases, tuition and expenses per student

Moving to Table 6, we see in Model 1 that factors ostensibly associated with faculty resistance did not operate as expected. Our proxy for the presence of a union was not significant and % full professors is associated with planning in the opposite direction of the one expected. Once other variables are included in Model 5, the effect of percentage full professors is eliminated. In Model 2, relationships with indicators of financial health are not all associated with the dependent variable in expected ways. Schools with higher endowment per student and higher net revenue (i.e. larger surplus) are *more* likely to return to in-person instruction.

**Table 6:**
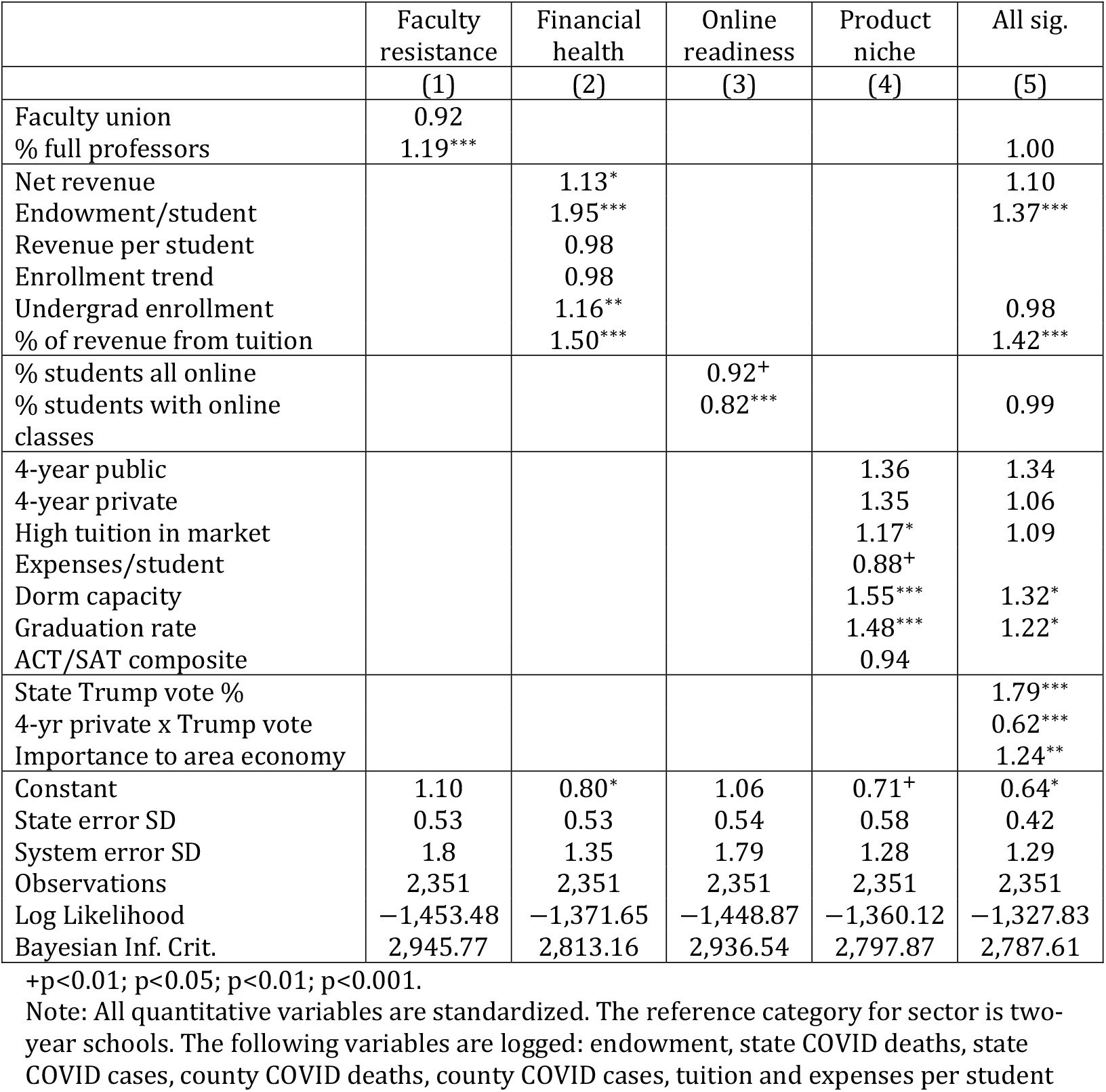
Hierarchical (two-level) logistic models examining August 1, 2020 college plans to have any in-person instruction in the fall 2020 semester cont.

Online readiness indicators in Model 3 both predict plans in expected ways. And the importance of product niche is evident from Model 4 of Table 6. In a series of regressions not shown, we found that the large differences across institutional types (two-year, four-year public and four-year private) that we saw in Table 4 are largely explained by variation in dorm capacity. We also see that institutions charging high tuitions within their market niche and those with high graduation rates were more likely to plan in-person classes.

In the final analysis, as shown in Model 5 of Table 6, we see that state Trump vote percentage matters most for public institutions. Other predictive factors include the percentage of revenue from tuition, endowment per student, the importance to area economy, dorm capacity and graduation rate.

## DISCUSSION AND CONCLUSION

This study focused on identifying the key predictors of college administrators’ decisions to return to in-person instruction as of August 1, 2020. We examined a wide range of factors that could have made a difference and were surprised to find that neither the proportion of COVID-19 cases, deaths, nor mask mandates had any association with colleges’ decisions to open their campuses. For public two- and four-year institutions, the proportion of state residents who voted for Trump was the most powerful predictor. Meanwhile, private institutions were relatively impervious to these political forces.

Public institutions in our sample received an average of about 39% of their core revenue from state government. For this reason, colleges located in places with a stronger pro-Trump orientation may have been under particularly strong pressure to fully reopen, even in light of COVID-19 infection rates and deaths (Desrochers and Hurlburt 2016). Conversely, likely because they do not rely on state funding, private colleges were less affected by this political element.

In our study we examined the role of tuition relative to college sector (i.e., public, private). We did this because sector is so highly correlated with tuition. While over four-fifths of private colleges charge over $20,000 a year, only one public college in the US is that expensive. We found that colleges with higher tuitions within their sector were indeed more likely to offer in-person instruction. Likewise, if colleges received more of their revenue from tuition, then they were also more likely to offer in-person instruction, as were those with higher graduation rates. Many colleges had to seriously consider the extent to which students would be willing to enroll if they could not offer a bespoke experience that many have come to expect (Sun 2020). While some of the most elite colleges, including Harvard and Princeton, announced before August that they would not be offering in-person instruction, other colleges could not be as confident that students would still attend or that they could weather the financial hardship if substantial numbers did not enroll.

We also found that colleges with larger endowments were more likely to offer in-person instruction. On the one hand, these colleges should have been better equipped to manage the financial challenges of potential lost enrollment. But, colleges with larger endowments are also more likely to provide a bespoke experience and offer activities and events (e.g., college football), that can provide a lot of additional revenue (Ilana 2020). Additionally, colleges with larger endowments may be better able to manage the costs associated with reopening (e.g., testing, cleaning, ventilation systems, etc.) (ACE 2020; CDC 2020a), especially given the potential of lost tuition dollars if they do not.

Finally, we found that the college’s importance to the local economy was a substantial predictor of reopening. Communities that are relatively small compared to the college population are financially dependent on students returning (Sullivan 2020). In addition to students spending money on housing, food, and entertainment, college towns are likely to draw people into the community for a wide range of amenities and activities, especially sporting and cultural activities, which fuel local businesses and tax dollars (Grady 2017). In deciding whether or not to reopen, these colleges would have had to consider the economic effects of not bringing students to campus, as well as the potential community backlash.

While we were able to identify several substantial predictors, many expected associations were negligible. Neither faculty preferences nor online readiness were ultimately associated with reopening decisions in a multivariate context in the final model. For explaining unique variation in opening decisions, money and politics seemed to matter more than anything with the state proportion who voted for Trump having the biggest effect size.

Our study provides an ‘aerial view’ of the subject matter. We did not talk with college administrators about their decision-making process and whether they considered, for example, the extent to which the surrounding area supported Trump’s election. One strength of our study is that they too may not have realized the role that such factors as state politics ultimately played. At the same time, there may have been other forces not examined here that had an important influence. Future research might consider conducting interviews to get a better sense of administrators’ perspectives.

Our study assessed a wide range of characteristics. To the best of our ability we used the most reliable measures we could find for measuring various concepts. But we did not always have a perfect match. For example, we thought that colleges with an older faculty population would advocate more for online classes, but we did not have a measure of average faculty age. Rather we relied on the college’s proportion of full faculty, which assesses the potential power of more senior faculty, but only inadvertently considers age.

Our study was focused on opening decisions as of August 1, 2020. We do not assess what colleges ultimately decided. However, their decision to fully reopen in-person as of August 1, 2020 is perhaps a better measure of preferences than what ultimately happened. As colleges began to provide in-person instruction in August and September 2020, many had to make emergency decisions to move temporarily or permanently online because of high numbers of on-campus COVID-19 cases (Burke 2020). Indeed, early reports suggested that college reopenings were playing a role in keeping US COVID-19 rates high (Abbott 2020; Hubler 2020). Understanding colleges’ intentions may provide insight into the factors shaping their decision making than what they ultimately did, where they may have had less control.

COVID-19 dramatically raised the stakes of decision-making by compelling colleges to engage in a cost-benefit analysis weighing the risks to health of individuals with that of their institutions. The severity of the local experience with COVID-19 appears to have played no role in these decisions. In part this is likely because at the time when decisions were made, administrators would have accurately ascertained that the risks of new infection rates at the time of school reopening might well be unrelated to cumulative infections in June or July when many reopening decisions were made. Perhaps this null result points as well to the fact COVID-19 had spread sufficiently by mid-summer that the risk of infection was perceived as having roughly the same order of magnitude nationwide, at least on college campuses.

Geographic variation in public perceptions of risk is unlikely itself to be a major factor here since state politics only affected public institutions. The fact that state political climate (measured by the state percentage voting for Trump in the 2016 election) affected public yet not private institutions strongly suggests instead that political influence on colleges occurred exclusively through the state education bureaucracy.

While college executives certainly considered many of the factors examined here, we likely captured some unanticipated pressures that were not overtly considered, suggesting that the decision-making process may be even less bounded by rationality than anticipated (i.e., Simon 1990). Given that the pandemic is likely to continue wreaking havoc for the foreseeable future, many colleges will have to make more key opening decisions. Some of the dynamics in our study are likely to operate in future decision making, as well as what colleges previously decided and what they see at other institutions.

## Data Availability

All data are available in the public domain. See article for information on where to get specific data.

## Footnotes

1 Our study excludes specialized schools (e.g., Yeshiva, art, engineering, culinary schools) and those where 50% or more of students are fully online, as we did not have good coverage in our sample.

2 Colleges in the United States typically offer mostly in-person instruction. Relatively few (e.g., culinary schools) provide all in-person instruction, or only online classes (e.g., Arizona).

3 In early September 2020, Insider Higher Education magazine noted that the College Crisis Initiative at Davidson College had conducted an analysis with reopening data, finding that the proportion of an area voting for Trump was associated with a greater likelihood of providing in-person instruction (Madeline 2020). However, the magazine did not provide much additional information, like the analysis technique or additional variables/controls examined, and did not mention almost any of the other factors that we consider. We tried to get more information about the study, but did not get a response. A thorough literature search did not find any related articles.

4 Two additional alternatives were (1) treating unaffiliated institutions as their own clusters in a random effects Model and (2) generalized estimating equations. The former encountered singularity problems in estimation and the latter was not equipped to handle the partially nested, cross-classified nature of these data in which one had to account for clustering within state *and* higher education systems.

